# The Association of Socioeconomic Status, the Concern for Catching Covid-19, and Anxiety Between Individuals with and without a Cancer History from a Cross-sectional Study

**DOI:** 10.1101/2022.07.26.22278080

**Authors:** Xiaochen Zhang, Sonya Sasmal, Mengda Yu, Brittany Bernardo, Toyin Adeyanju, Electra D. Paskett

## Abstract

**Background:** COVID-19 has negative impacts on mental health in all populations. Individuals with a history of cancer have an increased risk of catching and having more severe symptoms of COVID-19 than the general public. The objective of this study was to examine how cancer history and concern for catching COVID-19 relate to anxiety.

**Methods:** This cross-sectional study is part of the “Impact of COVID-19 on Behaviors across the Cancer Control Continuum in Ohio” project conducted from June to November 2020. The sample consisted of 7012 participants who completed survey online, by phone, or by mail. Self-reported concern for catching COVID-19 and anxiety over the last 7 days were used. Linear and logistic regression models were performed to determine the association between demographics, cancer history, concern for catching COVID-19, and anxiety.

**Results:** In our study sample, most participants rated their concern for catching COVID-19 as moderately high or high (56%) and reported anxiety for one day or more (63%). Individuals with a cancer history were more likely to report moderate-high or high concern for catching COVID-19 (59% vs.54%, P<0.001) but less likely to report anxiety (58% vs. 67%, P<0.001) compared to those without a cancer history. Individuals with higher SES were less likely to report anxiety (middle vs. low SES: OR=0.68, 95%CI=0.59-0.79; high vs. low SES: OR=0.70, 95%CI=0.61-0.82). Additionally, increased concern for catching COVID-19 was associated with higher likelihood of reporting anxiety (moderate-low vs. low: OR=1.65, 95%CI=1.42-1.92; moderate-high vs. low: OR=2.98, 95%CI=2.53-3.50; high vs. low: OR=4.35, 95%CI=3.74-5.07).

**Conclusions:** Our findings suggest individuals with a cancer history reported higher concern for catching COVID-19. Higher concern for catching COVID was associated with anxiety. These findings indicate that healthcare providers should pay special attention to the different populations to reduce concerns for catching COVID-19 and provide strategies to improve mental health during a pandemic outbreak.

**Funding:** This study was supported by a supplement to The Ohio State University Comprehensive Cancer Center (OSUCCC) core support grant (P30 CA016058), and the OSUCCC The Recruitment, Intervention and Survey Shared Resource (RISSR)(P30 CA016058).The Ohio State University Center for Clinical and Translational Science grant support (National Center for Advancing Translational Sciences, Grant UL1TR001070) in publications relating to this project. This work was supported by the National Cancer Institute (F99CA253745 to X.Z.).

## INTRODUCTION

Since March 2020, coronavirus disease 2019 (COVID-19) has spread with over 79 million individuals infected and 974,000 deaths in the United States (US) ^1^. Person to person transmission of the virus is spread through close contact and respiratory droplets when a person coughs and sneezes ^2^. Due to the highly infectious nature of the virus, widespread physical distancing measures, like stay-at-home orders, were put in place to decrease viral spread ^3^. Individuals with older age and pre-existing health conditions (e.g., obesity, heart disease, chronic kidney disease, cancer) are at risk for severe disease and death from COVID-19 ^4-6^. The COVID-19 pandemic has negative consequences beyond the virus itself ^7^. The overwhelming amount of information about the disease, concerns about morbidity, mortality, and long-term side effects, as well as changes in daily living were associated with psychological distress, anxiety, and depression ^3,7-12^. The fear of and concern about COVID-19 was linked to increased anxiety and depression levels ^13-15^.

Although the ongoing pandemic causes unfavorable impacts on mental health among all populations, COVID-19 has affected communities differently ^16-20^. For example, the economic and social stresses of job loss have fallen most heavily on individuals with lower socioeconomic status (SES). Individuals with low SES tend to have less ability to work from home during the pandemic, minimal to no health insurance, less access to healthcare, and lower health literacy, all of which can explain the high disease burden of COVID-19 in this population ^21^. Additionally, SES, chronic illness, and being in an at-risk group were positively associated with fear of COVID-19 ^15^. These effects may cause a psychological toll on individuals with low SES, and thus exacerbate pre-existing health disparities ^22,23^.

Individuals with a cancer history are another particular population that are at higher risk of developing severe symptoms, being hospitalized, admitted to the ICU, and dying from COVID-19 ^24-26^. To avoid contracting COVID-19 in health care settings, individuals with a cancer history had to delay or limit the frequency of clinical visits, including diagnosis, cancer treatment (e.g., surgery, chemotherapy, radiation), and palliative care, all of which are crucial to cancer prognosis ^24,27-30^. Moreover, individuals with a cancer history have a higher prevalence of anxiety and emotional distress compared to the general population before the COVID-19 pandemic ^31^. During the COVID-19 pandemic, in addition to the fear of COVID-19, individuals with a cancer history may feel even more overwhelmed, stressed, and worried about their cancer management, which lead to higher levels of psychological distress, depression, and anxiety ^32-40^.

Our study examined how SES and cancer history related to concern for catching COVID-19 and anxiety. The findings of this study can inform strategies and programs for vulnerable populations, such as individuals with low SES or a cancer history to address concerns of catching COVID and reduce anxiety.

## RESULTS

A total of 32989 individuals were invited and 9423 completed the survey (Figure 1). After removal of missing variables of interest (n=2411), a sample size of 7012 participants were included for the current analysis. Compared to individuals included in the analysis, individuals who were excluded were more likely to be older, male, and have missing SES (Supplementary Table 1, all p<0.001).

**Figure 1.**
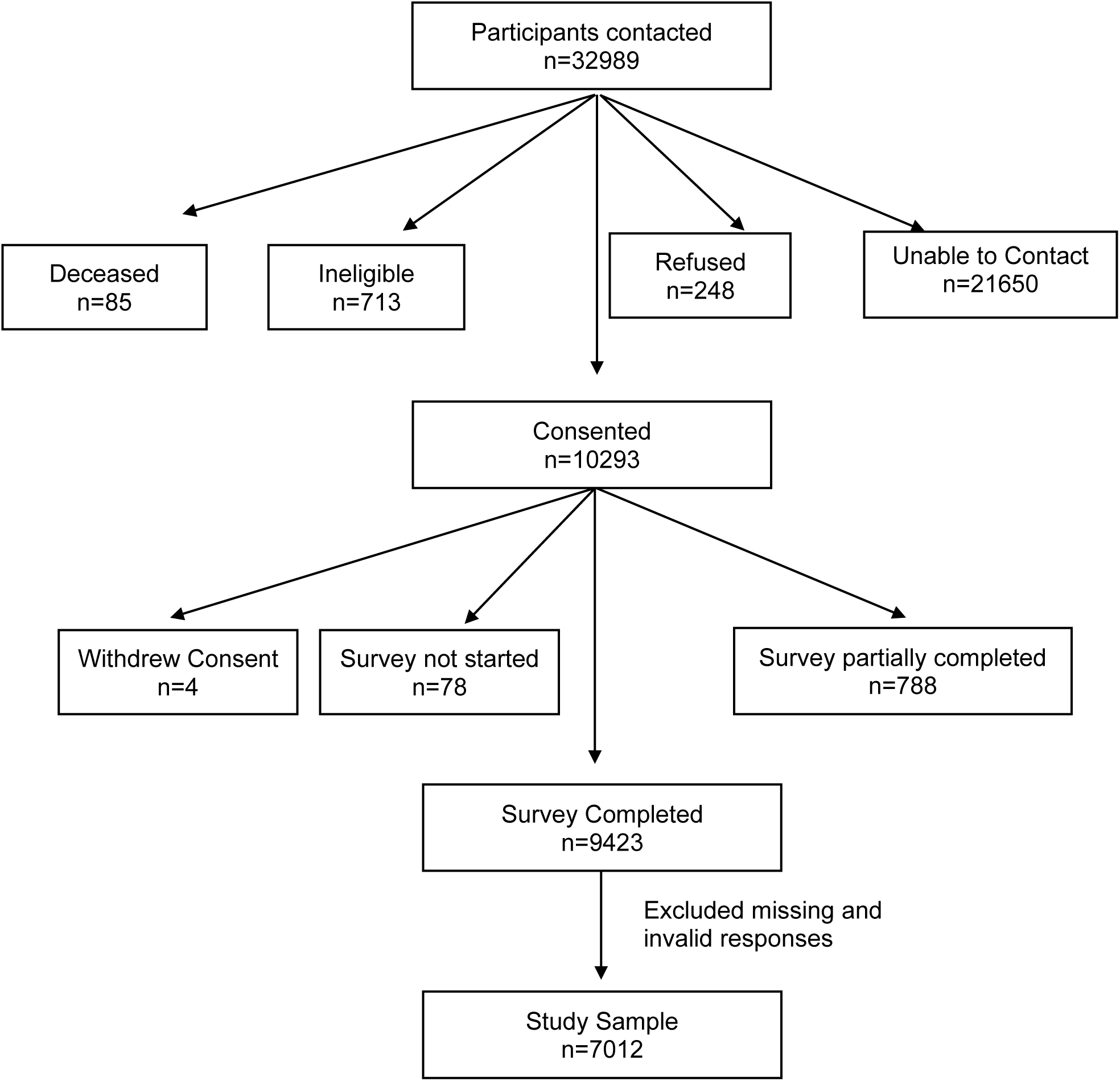
COVID-19 Survey Recruitment Diagram.

Among the included participants (n=7012), 49.2% had a history of cancer (n=3450) and 50.8% did not have a history of cancer (n=3562) (Table 1). The average age was 56.6 years, 32.2% were male, 89.6% were White, and 22.8% with low SES (SES score: 0-5), 41.7% with intermediate SES (SES score: 6-8), and 35.4% with high SES (SES score: 9-10). Compared to those with no history of cancer diagnosis, participants who had history of cancer diagnosis were older, more likely to be male, White, and had intermediate SES (Table 1; all p<0.05).

**Table 1.** Demographic characteristics of participants, overall and by history of cancer diagnosis.

| Variable | Total<br>N=7012 | No history of<br>cancer diagnosis<br>n=3562 (50.8%) | Had history of<br>cancer diagnosis<br>n=3450 (49.2%) | P-value |
| --- | --- | --- | --- | --- |
| Age, years, Mean±SD | 56.59±13.32 | 53.01±13.25 | 60.29±12.33 | <0.001 |
| ≤40 yrs | 997 (14.22%) | 724 (20.33%) | 273 (7.91%) |  |
| 41-60 yrs | 3012 (42.95%) | 1735 (48.71%) | 1277 (37.01%) | <.0001 |
| ≥61 yrs | 3003 (42.83%) | 1103 (30.97%) | 1900 (55.07%) |  |
| Sex |  |  |  |  |
| Male | 2260 (32.23%) | 916 (25.72%) | 1344 (38.96%) |  |
| Female | 4752 (67.77%) | 2646 (74.28%) | 2106 (61.04%) | <.0001 |
| Race |  |  |  |  |
| White | 6279 (89.55%) | 3150 (88.43%) | 3129 (90.7%) |  |
| Black | 412 (5.88%) | 227 (6.37%) | 185 (5.36%) |  |
| Asian | 122 (1.74%) | 62 (1.74%) | 60 (1.74%) | 0.0033 |
| Other/Multiple | 199 (2.84%) | 123 (3.45%) | 76 (2.2%) |  |
| SES score |  |  |  |  |
| Low (0-5) | 1601 (22.83%) | 795 (22.32%) | 806 (23.36%) |  |
| Intermediate (6-8) | 2929 (41.77%) | 1409 (39.56%) | 1520 (44.06%) | <.0001 |
| High (9-10) | 2482 (35.4%) | 1358 (38.12%) | 1124 (32.58%) |  |

### Concern of Catching COVID-19

Among all participants, 1369 (19.5%) reported low concern for catching COVID-19 (scale: 0-25) (Table 2), 1710 (24.4%) reported moderate to low concern (scale: 26-50), 1519 (21.7%) reported moderate to high concern (scale: 51-75), and 2414 (34.4%) reported high concern of catching COVID-19 (scale: 75-100). Compared to individuals with no history of cancer diagnosis, individuals that had a history of cancer diagnosis were more likely to have high concern (37.7% vs. 31.3%) and less likely to report low concern of catching COVID-19 (17.5% vs. 21.5%, p<0.0001).

**Table 2.** Concern of catching COVID and anxiety in the past 7 days, overall and by history of cancer diagnosis.

| Variable | Total<br>N=7012 | No history of cancer<br>diagnosis<br>n=3562 (50.8%) | Had history of<br>cancer diagnosis<br>n=3450 (49.2%) | P-value |
| --- | --- | --- | --- | --- |
| Concern of catching COVID |  |  |  |  |
| Low (0-25) | 1369 (19.52%) | 766 (21.5%) | 603 (17.48%) | <.0001 |
| Moderate low (26-50) | 1710 (24.39%) | 884 (24.82%) | 826 (23.94%) |  |
| Moderate high (51-75) | 1519 (21.66%) | 799 (22.43%) | 720 (20.87%) |  |
| High (75-100) | 2414 (34.43%) | 1113 (31.25%) | 1301 (37.71%) |  |
| Anxiety in the past 7 days |  |  |  |  |
| Not at all | 2600 (37.08%) | 1164 (32.68%) | 1436 (41.62%) | <.0001 |
| One day or more | 4412 (62.92%) | 2398 (67.32%) | 2014 (58.38%) |  |

When examining the association of demographic characteristics, history of cancer diagnosis, and concern of catching COVID-19, the relative risk of reporting moderate low concern vs. low concern was higher among female (Table 3, OR=1.46, 95% CI=1.25-1.71), Asian (OR=2.69, 95% CI=1.31-5.56), intermediate SES (OR=1.26, 95% CI=1.06-1.52), high SES (OR=1.26, 95% CI=1.03-1.53), and individuals with history of cancer diagnosis (OR=1.21, 95% CI=1.04-1.40). The relative risk of reporting moderate high concern vs. low concern was lower among those who were 41-60 years old (OR=0.63, 95% CI=0.50-0.78), but higher among female (OR=1.38, 95% CI=1.18-1.62), Black (OR=1.60, 95% CI=1.14-2.24), intermediate SES (OR=1.36, 95% CI=1.11-1.66), high SES (OR=1.84, 95% CI=1.50-2.27), and individuals with history of cancer diagnosis (OR=1.17, 95% CI=1.00-1.37). Similarly, the relative risk of reporting moderate high concern vs. low concern was lower among those who were 41-60 years old (OR=0.66, 95% CI=0.54-0.82) and higher among female (OR=1.35, 95% CI=1.17-1.57) and individuals with history of cancer diagnosis (OR=1.50, 95% CI=1.30-1.73). However, compared to White, Black (OR=1.85, 95% CI=1.36-2.50), Asian (OR=4.11, 95% CI=2.08-8.09), and Other race (OR=1.50, 95% CI=1.01-1.35) individuals had a higher relative risk of reporting high concern vs. low concern for catching COVID-19.

**Table 3.**
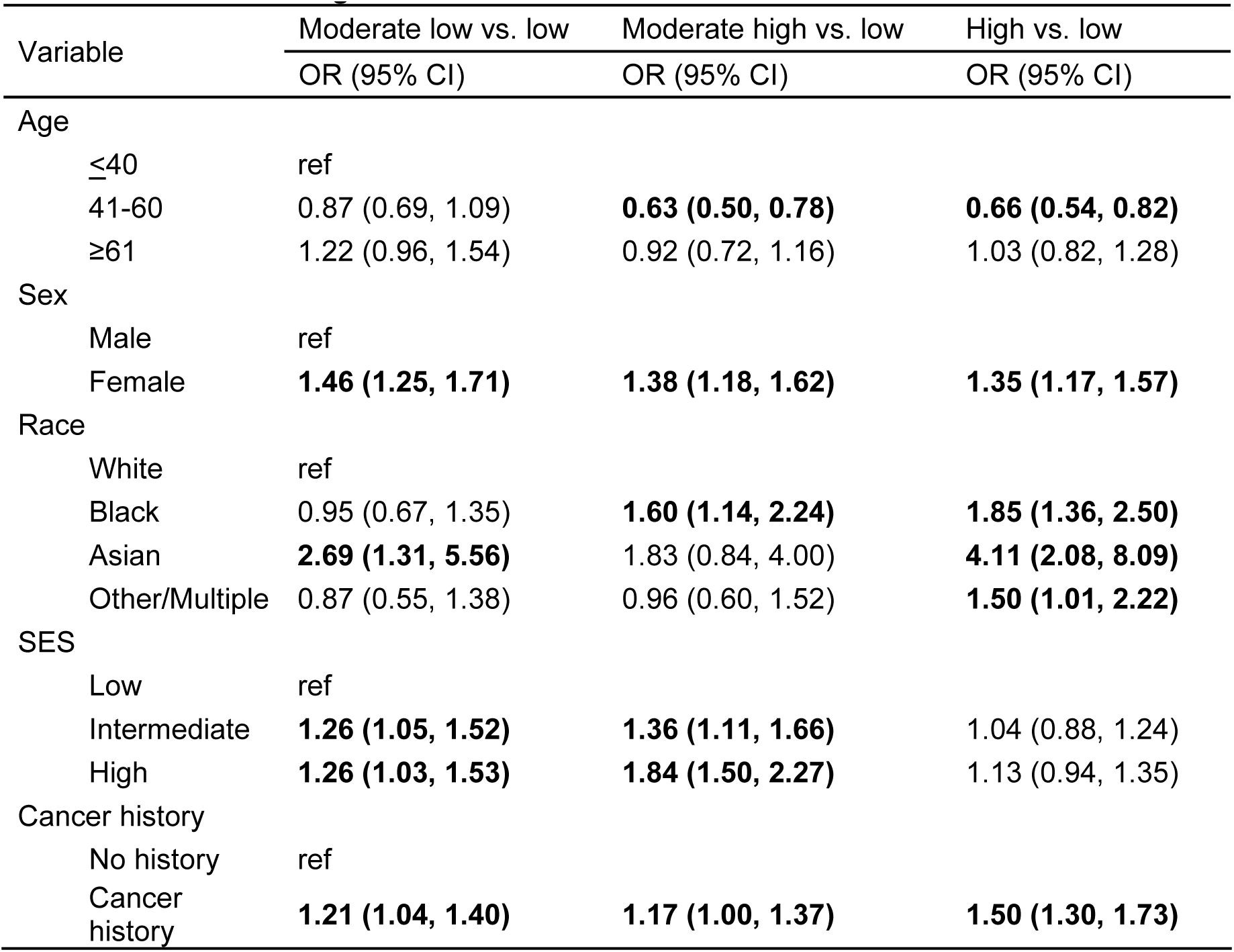
The association of demographic characteristics, history of cancer diagnosis, and concern for catching COVID.

### Anxiety

Among all participants, 2600 (37.1%) reported no anxiety in the past 7 days (Table 2), and 4412 (62.9%) reported they felt nervous, anxious, or on edge one day or more in the past 7 days. Interestingly, compared to individuals with no history of cancer diagnosis, individuals who had a history of cancer diagnosis were less likely to have anxiety in the past 7 days (58.4% vs. 67.3%, p<0.0001).

After adjusting for history of cancer diagnosis, participants who were 41-60 years and ≥61 years had lower odds of reporting anxiety, compared to participants ≤40 years old (Table 4, model 1; OR=0.43, 95% CI=0.36-0.52; OR=0.22, 95% CI=0.18-0.26, respectively). Asian participants had lower odds of reporting anxiety vs. White participants (OR=0.54, 95% CI=0.37-0.79). Participants with intermediate (OR=0.71, 95% CI=0.62-0.81) and high SES (OR=0.76, 95% CI=0.66-0.87) also had lower odds of reporting anxiety vs. participants with low SES. Female participants had higher odds of reporting anxiety compared to male participants (OR=1.92, 95% CI=1.72-2.14). Similar results were observed between demographic characteristics and anxiety after adjusting for concern of catching COVID-19 (Table 4, model 2). Interestingly, those who had history of cancer diagnosis had lower odds of anxiety. However, higher level of concern for catching COVID-19 were associated with increased odds of anxiety (moderate low vs. low: OR=1.65, 95% CI=1.42-1.92; moderate high vs. low: OR=2.98, 95% CI=2.53-3.50; high vs. low: OR=4.35, 95% CI=3.74-5.07).

**Table 4.** The association of demographic characteristics, history of cancer diagnosis, concern of catching COVID-19, and anxiety.

| Variable | Model 1<br>OR (95% CI) | Model 2<br>OR (95% CI) |
| --- | --- | --- |
| Age |  |  |
| ≤40 | ref |  |
| 41-60 | 0.43 (0.36, 0.52) | 0.45 (0.37, 0.55) |
| ≥61 | 0.22 (0.18, 0.26) | 0.20 (0.16, 0.24) |
| Sex |  |  |
| Male | ref |  |
| Female | 1.92 (1.72, 2.14) | 1.91 (1.71, 2.13) |
| Race |  |  |
| White | ref |  |
| Black/Afr American | 0.97 (0.78, 1.22) | 0.84 (0.66, 1.06) |
| Asian | 0.54 (0.37, 0.79) | 0.42 (0.28, 0.62) |
| Other/Multiple | 1.07 (0.77, 1.49) | 0.99 (0.70, 1.40) |
| SES |  |  |
| Low | ref |  |
| Middle | 0.71 (0.62, 0.81) | 0.68 (0.59, 0.79) |
| High | 0.76 (0.66, 0.87) | 0.70 (0.61, 0.82) |
| Cancer history |  |  |
| No history | ref |  |
| Cancer history | 0.95 (0.86, 1.06) | 0.87 (0.78, 0.97) |
| Concern of catching COVID |  |  |
| Low | -- | ref |
| Moderate low | -- | 1.65 (1.42, 1.92) |
| Moderate high | -- | 2.98 (2.53, 3.50) |
| High | -- | 4.35 (3.74, 5.07) |

Model 1: adjusted for age, sex, race, SES, cancer history
Model 2: adjusted for age, sex, race, SES, cancer history AND concern of catching COVID-19

## DISCUSSION

Our study showed that most participants rated their concern for catching COVID-19 as high or moderate high, however, a larger proportion of individuals with a history of cancer fell within the high concern group compared to individuals without a history of cancer. Since the survey responses were received during the height of the pandemic, between June 2020 and November 2020, these findings are not surprising, as individuals with a cancer diagnosis are at higher risk for developing complications from COVID-19. ^(1,4,10)^. Our finding aligns with the hypothesis that individuals with a cancer history report higher concern for catching COVID-19.

Among individuals with and without a history of cancer, the majority were classified within the middle SES group/level. While there was no association between SES and high concern for catching COVID-19, we observed that participants in higher SES levels were more likely to report moderate-low and moderately-high concern for catching COVID-19 than those with a low SES level. When examined the association between SES and anxiety, we found that individuals with middle or high SES were less likely to report anxiety compared to those with low SES, which was consistent with our hypothesis.

We hypothesized that individuals with a cancer history would be more likely to report anxiety than individuals without cancer history. Interestingly, we observed that individuals with a cancer history were actually less likely to report anxiety compared to those without a cancer history. This finding differed from previous study by Niedzwiedz et. al, which indicates higher levels of anxiety experienced by cancer patients when compared to the general population ^31^. This could be explained that the anxiety assessment used in our study differed from other validated measures. After controlling for age, sex, race, and SES, individuals with and without a history of cancer had similar odds of reporting anxiety. This is consistent with the findings from van de Poll-Franse et. al, that observed no differences in anxiety, depression, and quality of life between cancer patients and non-cancer participants during the COVID-19 pandemic ^11^.

Our results demonstrated that the increased level of concern for catching COVID-19 was associated with increased likelihood of reporting anxiety. This is consistent with established links between pandemic-related anxiety and elevated concerns of catching diseases in various global disease outbreaks ^41-43^. We also found some notable differences in anxiety reported by certain populations, after controlling for the level of concern for catching COVID-19. Individuals with older age, middle or high SES, a cancer history, or who were Asian were less likely to report anxiety, compared to their counterparts. However, female participants were more likely to report anxiety, which is consistent with previous findings that females have a higher rate of anxiety and psychological distress compared with males ^44-47^. These findings suggested that resources and programs to address the impact of COVID on psychological health are needed, especially for women and individuals with low SES.

Strengths of this study include the large sample size of individuals with (n=3450) and without a history of cancer (n=3562). This allowed us to compare the associations between those with and without a cancer history in adults of all races, ages, and SES levels. Additionally, the timing of this study during the pandemic enabled us to get real-time data of the impact of COVID-19.

Limitations include the way anxiety was assessed. Our study used a single-item question to assess the number of days that participant experienced on edge, nervous, or anxious in the last 7 days. Using a more validated approach to assess anxiety, such as the COVID-19 Anxiety Scale (CAS), could allow us comparing findings across studies ^48,49^. In addition, we recruited participants from previous studies. It is possible that the impact of COVID-19 differs between individuals who participated vs. did not participate in research studies. Thus, our findings may not be generalizable to other populations.

We examined the associations of SES, concern for catching COVID-19, and anxiety between individuals with and without a cancer history. Our findings demonstrated higher concern for catching COVID among individuals with a cancer history, and high prevalence of anxiety regardless cancer history in the early COVID pandemic (June to November 2020). Healthcare providers, policymakers, and other stakeholders should pay special attention to address concern of COVID, perhaps providing care and resources for those who are in need, to improve mental health.

## MATERIALS AND METHODS

### Overview

This study was part of an NCI-funded initiative conducted in conjunction with 16 other NCI-designated Cancer Centers, the IC-4 (Impact of COVID-19 on the Cancer Continuum Consortium). The initiative was funded to work collectively to develop core survey items and implement population surveys in the respective catchment areas. The overall goal of the IC-4 was to assess how differences in demographics (rural/urban, age, gender, race, educational attainment) impact engagement in cancer preventive behaviors (e.g., tobacco cessation, screening, diet) and cancer management/ survivorship behaviors (e.g., adherence to treatment, adherence to surveillance, access to health services) in the context of COVID-19 environmental constraints (e.g., social distancing, employment, mental health). Each site had its own theoretical framework and survey methods. Our site used the IC-4 core set of common data elements, with remote data collection methods to include many unique and diverse populations. This study was approved by the OSU Institutional Review Board in June 2020.

### Theoretical Framework

This study was grounded in the Health Belief Model (HBM) ^50,51^. According to the HBM, individuals’ change in health behaviors depends on a series of health beliefs, include: 1) perceived susceptibility to COVID-19 exposure, 2) perceived severity of the consequences of contracting COVID-19 (e.g., hospitalization or death), 3) perceived benefits of the effectiveness of the proposed COVID-19 prevention measures, 4) perceived barriers to executing the proposed prevention measures, 5) cue to the proposed prevention actions, and 6) self-efficacy in the person’s ability to successfully perform COVID-19 prevention measures.

### Survey Development

The survey elements (See Appendix 1) were finalized in conjunction with other members of the IC-4^52^. The survey included individual behaviors related to mitigation of COVID-19 transmission, the challenges related to social distancing, self/family isolation, stress, and health behaviors that are highly relevant to cancer and other chronic diseases (i.e., type, duration and location of physical activity, tobacco/marijuana or alcohol use, vaping/e-cig use, exposure to second hand smoke, nutrition/diet, health information-seeking and participation in clinical trials, and access to health services). Questions also assessed perceived stigma related to COVID-19 with respect to different population groups and covariates, such as health literacy and mental health, suspected of moderating these influences. Moreover, we assessed the impact of children being out of school and employment challenges (i.e. remote working and unemployment etc.), as well as the influence of social media on information, knowledge, behaviors and attitudes.

### Sample Selection

Eligible participants were adults aged 18 years or older, including healthy adult volunteers, cancer patients, cancer survivors, and cancer patients and survivors’ caregivers, mostly from Ohio, some from Indiana and other states. Two recruitment strategies were employed at OSU. First, we identified and contacted individuals who previously participated in studies and agreed to be contacted for future research projects. We also invited cancer patients and survivors to nominate their primary caregivers to participate in the study. The list of previous research projects conducted at OSU included the Rural Interventions for Screening Effectiveness (RISE) study (R01 CA196243), the Community Initiative Towards Improving Equity and Health Status (CITIES) cohort (Supplement to P30CA016058), the Buckeye Teen Health Study (BTHS) study (P50CA180908), the Ohio State University Center of Excellence in Regulatory Tobacco Science (CERTS) cohort (P50CA180908), and members of the Total Cancer Care (TCC) cohort (P30CA016058). Second, to further enhance the representativeness of our study sample and ensure the inclusion of minority and underserved communities, we utilized our community partners and listservs to send tailored email invitations.

### Interview/Data Collection

We utilized several data collection methods, including web, phone, and mailed surveys. Respondents with valid emails received an initial survey invitation email along with three reminders seven days apart. All participants were initially screened using an eligibility form before conducting the survey. Participants were able to save the web survey and resume it at a later time. Those who partially completed the web survey received an email reminder one week after they last accessed the survey. A trained interviewer contacted participants without an email address and those with invalid emails on file by phone. Participants who were initially reached by phone were offered the option to complete the survey over the phone or online. We mailed a cover letter and a paper survey with a self-addressed, stamped return envelope to participants who requested a mailed survey. For Non-English-speaking participants, a bilingual staff member administered the survey in the appropriate language. Participants were offered a $10 gift card upon completion of the survey. All data were collected and managed using the Research Electronic Data Capture (REDCap) secure web-based application hosted at OSU. This study followed the International Committee of Medical Journal Editors (ICMJE) guideline. The CONSORT diagram (**Figure 1**) demonstrated study enrollment.

### Measurement

To assess concern for catching COVID-19, participants were asked “From 0 to 100, how concerned are you about catching COVID-19? 0=Not at all concerned; 100=Extremely concerned.” Participants were classified as low (0-25), moderately low (26-50), moderately high (51-75), and high (76-100) concern. The main outcome of this study, anxiety, was assessed as, “In the past 7 days, have you felt nervous, anxious, or on edge?” Participants were classified as “not at all” or “one day or more”.

In terms of history of cancer diagnosis, participants were asked whether a doctor has ever diagnosed them with cancer. A modified Hollingshead Score was used to define socioeconomic status (SES) from insurance status, occupation, education level, and household income ^53^. SES for each participant was scored on a 0-10 scale by summing the individual scores for the four variables listed above and grouped into three groups: low SES (score: 0-5), intermediate SES (score: 6-8), and high SES (score: 9-10). Other demographic variables, such as age, race, and sex were included in the statistical analysis.

### Statistical Analysis

Characteristics were summarized using descriptive statistics including means and standard deviations for the continuous variables and frequencies for the categorical variables overall, and by history of cancer diagnosis. Differences between those who had cancer diagnosis vs. no history of cancer diagnosis were compared using two-sample T test or Kruskal-Wallis test for the continuous variables and Chi-square test or Fisher’s exact test for the categorical variables.

We assessed the association of demographic characteristics, history of cancer diagnosis, and concern for catching COVID-19, multinomial logistic regression models were used to estimate the odds ratios and 95% confident interval of moderate low vs. low concern, moderate high vs. low concern, and high vs. low concern of catching COVID-19. To assess the association between demographic characteristics, history of cancer diagnosis, concern for catching COVID-19 and anxiety, multivariable logistic regression was used. All analysis performed using the SAS 9.4 software.

## Data Availability

The data are not publicly available, however, the data that support the findings of this study are available from the corresponding author upon reasonable request and approval of the project publication committee.

## DATA AVAILABILITY

This is an ongoing study with Human Subjects. De-identified data will be available to access once the study ends. Interested researchers can request the data that support the findings of this study by contacting the corresponding author. The procedure outlined below must be followed:

1. First, the researcher must submit a short proposal to the Project Publication Committee for approval. This should include study rationale, introduction, methods, aims, data/variables and hypothesis as well as dummy tables.
2. The Project Publication Committee will review the proposal and make suggestions and/or recommendations.
3. Once the proposal has been approved, the project statistician will start working on the analysis. The statistician and the investigators will then meet to discuss the paper and the analysis plans. The role of the statistician is to perform all analyses to ensure that methods are appropriate and statistically valid.
4. If it has been determined that the researcher will be performing the analysis (this needs to be requested in the initial proposal to the Project Publication Committee), then the researcher needs to complete the Data Distribution and Agreement Form to the Project Publication Committee. This form is a request for the specific data that the researcher needs as well as a notice of all policies and rules about using Impact of COVID-19 data.

## CONFLICT OF INTEREST

Electra Paskett would like to disclose that she has grant funding for work outside of this project from the Merck Foundation, Genentech and Pfizer. All other authors report there are no conflicts of interest.

**Supplementary Table 1.** Demographic characteristics for included and excluded participants.

| Variable |  | All responded participants<br>(n=9280) | Participants included<br>(n=7012) | Participants excluded<br>(n=2268) | P-value |
| --- | --- | --- | --- | --- | --- |
| Age |  |  |  |  |  |
|  | ≤40 yrs | 1349 (14.54%) | 997 (14.22%) | 352 (15.52%) | <.0001 |
|  | 41-60 yrs | 3786 (40.8%) | 3012 (42.95%) | 774 (34.13%) |  |
|  | ≥61 yrs | 4074 (43.9%) | 3003 (42.83%) | 1071 (47.22%) |  |
|  | Missing | 71 (0.77%) | 0 (0%) | 71 (3.13%) |  |
| Sex |  |  |  |  |  |
|  | Male | 3102 (33.43%) | 2260 (32.23%) | 842 (37.13%) | <.0001 |
|  | Female | 6130 (66.06%) | 4752 (67.77%) | 1378 (60.76%) |  |
|  | Missing | 48 (0.52%) | 0 (0%) | 48 (2.12%) |  |
| Race |  |  |  |  |  |
|  | White | 8201 (88.37%) | 6279 (89.55%) | 1922 (84.74%) | 0.1317 |
|  | Black | 559 (6.02%) | 412 (5.88%) | 147 (6.48%) |  |
|  | Asian | 162 (1.75%) | 122 (1.74%) | 40 (1.76%) |  |
|  | Other/Multiple | 245 (2.64%) | 199 (2.84%) | 46 (2.03%) |  |
|  | Missing | 113 (1.22%) | 0 (0%) | 113 (4.98%) |  |
| SES score |  |  |  |  |  |
|  | Low (0-5) | 1754 (18.9%) | 1601 (22.83%) | 153 (6.75%) | <.0001 |
|  | Intermediate (6-8) | 3081 (33.2%) | 2929 (41.77%) | 152 (6.7%) |  |
|  | High (9-10) | 2569 (27.68%) | 2482 (35.4%) | 87 (3.84%) |  |
|  | Missing | 1876 (20.22%) | 0 (0%) | 1876 (82.72%) |  |

## REFERENCES

1. CDC. Trends in number of COVID-19 cases in the US reported to CDC, by state/territory: daily trends in number of COVID-19 cases in the United States reported to CDC. Centers for disease Control and Prevention. COVID Data Tracker Web site. https://covid.cdc.gov/covid-data-tracker/#datatracker-home. Accessed 3/29, 2022.

2. Harapan H, Itoh N, Yufika A, et al. Coronavirus disease 2019 (COVID-19): A literature review. Journal of infection and public health. 2020;13(5):667–673.

3. Tull MT, Edmonds KA, Scamaldo KM, Richmond JR, Rose JP, Gratz KL. Psychological Outcomes Associated with Stay-at-Home Orders and the Perceived Impact of COVID-19 on Daily Life. Psychiatry Research. 2020;289:113098.

4. Estiri H, Strasser ZH, Klann JG, Naseri P, Wagholikar KB, Murphy SN. Predicting COVID-19 mortality with electronic medical records. NPJ digital medicine. 2021;4(1):1–10.

5. Yek C, Warner S, Wiltz JL, et al. Risk Factors for Severe COVID-19 Outcomes Among Persons Aged≥ 18 Years Who Completed a Primary COVID-19 Vaccination Series—465 Health Care Facilities, United States, December 2020– October 2021. Morbidity and Mortality Weekly Report. 2022;71(1):19.

6. Ho FK, Petermann-Rocha F, Gray SR, et al. Is older age associated with COVID-19 mortality in the absence of other risk factors? General population cohort study of 470,034 participants. PLoS One. 2020;15(11):e0241824.

7. Sønderskov KM, Dinesen PT, Santini ZI, Østergaard SD. The depressive state of Denmark during the COVID-19 pandemic. Acta Neuropsychiatr. 2020;32(4):226–228.

8. Wang C, Pan R, Wan X, et al. Immediate Psychological Responses and Associated Factors during the Initial Stage of the 2019 Coronavirus Disease (COVID-19) Epidemic among the General Population in China. Int J Environ Res Public Health. 2020;17(5).

9. Vindegaard N, Benros ME. COVID-19 pandemic and mental health consequences: Systematic review of the current evidence. Brain Behav Immun. 2020;89:531–542.

10. Torales J, O’Higgins M, Castaldelli-Maia JM, Ventriglio A. The outbreak of COVID-19 coronavirus and its impact on global mental health. Int J Soc Psychiatry. 2020;66(4):317–320.

11. van de Poll-Franse lv, de Rooij BH, Horevoorts NJE, et al. Perceived Care and Well-being of Patients With Cancer and Matched Norm Participants in the COVID-19 Crisis: Results of a Survey of Participants in the Dutch PROFILES Registry. JAMA Oncology. 2021;7(2):279–284.

12. Ornell F, Schuch JB, Sordi AO, Kessler FHP. “Pandemic fear” and COVID-19: mental health burden and strategies. In. Vol 42: SciELO Brasil; 2020:232–235.

13. Mistry SK, Ali A, Akther F, Yadav UN, Harris MF. Exploring fear of COVID-19 and its correlates among older adults in Bangladesh. Global Health. 2021;17(1):47.

14. Millroth P, Frey R. Fear and anxiety in the face of COVID-19: Negative dispositions towards risk and uncertainty as vulnerability factors. J Anxiety Disord. 2021;83:102454.

15. Tzur Bitan D, Grossman-Giron A, Bloch Y, Mayer Y, Shiffman N, Mendlovic S. Fear of COVID-19 scale: Psychometric characteristics, reliability and validity in the Israeli population. Psychiatry Res. 2020;289:113100.

16. Pashazadeh Kan F, Raoofi S, Rafiei S, et al. A systematic review of the prevalence of anxiety among the general population during the COVID-19 pandemic. J Affect Disord. 2021;293:391–398.

17. Batterham PJ, Calear AL, McCallum SM, et al. Trajectories of depression and anxiety symptoms during the COVID-19 pandemic in a representative Australian adult cohort. Med J Aust. 2021;214(10):462–468.

18. Robinson E, Sutin R. A, Jones, A, Daly, M, 2021. A systematic review and meta-analysis of longitudinal cohort studies comparing mental health before versus during the COVID-19 pandemic. medRxiv.

19. Brief P. COVID-19 and the Need for Action on Mental Health. World Health Organization. 2020.

20. Santomauro DF, Mantilla Herrera AM, Shadid J, et al. Global prevalence and burden of depressive and anxiety disorders in 204 countries and territories in 2020 due to the COVID-19 pandemic. The Lancet. 2021;398(10312):1700–1712.

21. Rozenfeld Y, Beam J, Maier H, et al. A model of disparities: risk factors associated with COVID-19 infection. International Journal for Equity in Health. 2020;19(1):126.

22. Patel V, Burns JK, Dhingra M, Tarver L, Kohrt BA, Lund C. Income inequality and depression: a systematic review and meta-analysis of the association and a scoping review of mechanisms. World Psychiatry. 2018;17(1):76–89.

23. Balogun OD, Bea VJ, Phillips E. Disparities in Cancer Outcomes Due to COVID-19—A Tale of 2 Cities. JAMA Oncology. 2020;6(10):1531–1532.

24. Gosain R, Abdou Y, Singh A, Rana N, Puzanov I, Ernstoff MS. COVID-19 and Cancer: a Comprehensive Review. Current Oncology Reports. 2020;22(5):53.

25. Schade EC, Elkaddoum R, Kourie HR. The psychological challenges for oncological patients in times of COVID-19 pandemic: telemedicine, a solution? Future oncology. 2020;16(29):2265–2268.

26. Sun L, Surya S, Le AN, et al. Rates of COVID-19–Related Outcomes in Cancer Compared With Noncancer Patients. JNCI Cancer Spectrum. 2021;5(1).

27. The Lancet O. COVID-19 and cancer: 1 year on. The Lancet Oncology. 2021;22(4):411.

28. Fillon M. Cancer treatment delays caused by the COVID-19 pandemic may not hinder outcomes. CA: A Cancer Journal for Clinicians. 2021;71(1):3–6.

29. Patt D, Gordan L, Diaz M, et al. Impact of COVID-19 on Cancer Care: How the Pandemic Is Delaying Cancer Diagnosis and Treatment for American Seniors. JCO Clinical Cancer Informatics. 2020(4):1059–1071.

30. Riera R, Bagattini ÂM, Pacheco RL, Pachito DV, Roitberg F, Ilbawi A. Delays and Disruptions in Cancer Health Care Due to COVID-19 Pandemic: Systematic Review. JCO Global Oncology. 2021(7):311–323.

31. Niedzwiedz CL, Knifton L, Robb KA, Katikireddi SV, Smith DJ. Depression and anxiety among people living with and beyond cancer: a growing clinical and research priority. BMC Cancer. 2019;19(1):943.

32. Yildirim OA, Poyraz K, Erdur E. Depression and anxiety in cancer patients before and during the SARS-CoV-2 pandemic: association with treatment delays. Qual Life Res. 2021;30(7):1903–1912.

33. Obispo-Portero B, Cruz-Castellanos P, Jiménez-Fonseca P, et al. Anxiety and depression in patients with advanced cancer during the COVID-19 pandemic. Support Care Cancer. 2022;30(4):3363–3370.

34. Gallagher S, Bennett KM, Roper L. Loneliness and depression in patients with cancer during COVID-19. J Psychosoc Oncol. 2021;39(3):445–451.

35. Massicotte V, Ivers H, Savard J. COVID-19 Pandemic Stressors and Psychological Symptoms in Breast Cancer Patients. Curr Oncol. 2021;28(1):294–300.

36. Miaskowski C, Paul SM, Snowberg K, et al. Stress and Symptom Burden in Oncology Patients During the COVID-19 Pandemic. J Pain Symptom Manage. 2020;60(5):e25–e34.

37. Moraliyage H, De Silva D, Ranasinghe W, et al. Cancer in Lockdown: Impact of the COVID-19 Pandemic on Patients with Cancer. Oncologist. 2021;26(2):e342–e344.

38. Ng KYY, Zhou S, Tan SH, et al. Understanding the Psychological Impact of COVID-19 Pandemic on Patients With Cancer, Their Caregivers, and Health Care Workers in Singapore. JCO Glob Oncol. 2020;6:1494–1509.

39. Rodrigues-Oliveira L, Kauark-Fontes E, Alves CGB, et al. COVID-19 impact on anxiety and depression in head and neck cancer patients: A cross-sectional study. Oral Dis. 2021.

40. Wang Y, Duan Z, Ma Z, et al. Epidemiology of mental health problems among patients with cancer during COVID-19 pandemic. Translational Psychiatry. 2020;10(1):263.

41. Chong MY, Wang WC, Hsieh WC, et al. Psychological impact of severe acute respiratory syndrome on health workers in a tertiary hospital. Br J Psychiatry. 2004;185:127–133.

42. Orrù G, Bertelloni D, Diolaiuti F, Conversano C, Ciacchini R, Gemignani A. A Psychometric Examination of the Coronavirus Anxiety Scale and the Fear of Coronavirus Disease 2019 Scale in the Italian Population. Frontiers in Psychology. 2021;12.

43. Wheaton MG, Abramowitz JS, Berman NC, Fabricant LE, Olatunji BO. Psychological predictors of anxiety in response to the H1N1 (swine flu) pandemic. Cognitive Therapy and Research. 2012;36(3):210–218.

44. Seo D, Ahluwalia A, Potenza MN, Sinha R. Gender differences in neural correlates of stress-induced anxiety. Journal of neuroscience research. 2017;95(1-2):115–125.

45. Leach LS, Christensen H, Mackinnon AJ, Windsor TD, Butterworth P. Gender differences in depression and anxiety across the adult lifespan: the role of psychosocial mediators. Social psychiatry and psychiatric epidemiology. 2008;43(12):983–998.

46. Kessler RC. Epidemiology of women and depression. Journal of affective disorders. 2003;74(1):5–13.

47. Bijl RV, De Graaf R, Ravelli A, Smit F, Vollebergh WA. Gender and age-specific first incidence of DSM-III-R psychiatric disorders in the general population. Results from the Netherlands Mental Health Survey and Incidence Study (NEMESIS). Soc Psychiatry Psychiatr Epidemiol. 2002;37(8):372–379.

48. Lee SA. Coronavirus Anxiety Scale: A brief mental health screener for COVID-19 related anxiety. Death Stud. 2020;44(7):393–401.

49. Silva WAD, de Sampaio Brito TR, Pereira CR. COVID-19 anxiety scale (CAS): Development and psychometric properties. Curr Psychol. 2020:1–10.

50. Janz NK, Becker MH. The Health Belief Model: a decade later. Health Educ Q. 1984;11(1):1–47.

51. Rosenstock IM. The Health Belief Model: explaining health behavior through experiences. Health behavior and health education: Theory, research and practice. 1990:39–63.

52. Scarinci IC, Pandya VN, Kim Y-i, et al. Factors associated with perceived susceptibility to COVID-19 among urban and rural adults in Alabama. Journal of Community Health. 2021:1–10.

53. Hollingshead AB. Four factor index of social status. In: New Haven, CT; 1975.

